# Increased travel times to United States SARS-CoV-2 testing sites: a spatial modeling study

**DOI:** 10.1101/2020.04.25.20074419

**Authors:** Benjamin Rader, Christina M. Astley, Karla Therese L. Sy, Kara Sewalk, Yulin Hswen, John S. Brownstein, Moritz U.G. Kraemer

## Abstract

**Importance:** Access to testing is key to a successful response to the COVID-19 pandemic.

**Objective:** To determine the geographic accessibility to SARS-CoV-2 testing sites in the United States, as quantified by travel time.

**Design:** Cross-sectional analysis of SARS-CoV-2 testing sites as of April 7, 2020 in relation to travel time.

**Setting:** United States COVID-19 pandemic.

**Participants:** The United States, including the 48 contiguous states and the District of Columbia.

**Exposures:** Population density, percent minority, percent uninsured, and median income by county from the 2018 American Community Survey demographic data.

**Main Outcome:** SARS-CoV-2 testing sites identified in two national databases (Carbon Health and CodersAgainstCovid), geocoded by address. Median county 1 km^2^ gridded friction surface of travel times, as a measure of geographic accessibility to SARS-CoV-2 testing sites.

**Results:** 6,236 unique SARS-CoV-2 testing sites in 3,108 United States counties were identified. Thirty percent of the U.S. population live in a county (N = 1,920) with a median travel time over 20 minutes. This was geographically heterogeneous; 86% of the Mountain division population versus 5% of the Middle Atlantic population lived in counties with median travel times over 20 min. Generalized Linear Models showed population density, percent minority, percent uninsured and median income were predictors of median travel time to testing sites. For example, higher percent uninsured was associated with longer travel time (*β* = 0.41 min/percent, 95% confidence interval 0.3-0.53, *p* = 1.2×10^−12^), adjusting for population density.

**Conclusions and Relevance:** Geographic accessibility to SARS-Cov-2 testing sites is reduced in counties with lower population density and higher percent of minority and uninsured, which are also risk factors for worse healthcare access and outcomes. Geographic barriers to SARS-Cov-2 testing may exacerbate health inequalities and bias county-specific transmission estimates. Geographic accessibility should be considered when planning the location of future testing sites and interpreting epidemiological data.

**Key Points:**

1. SARS-CoV-2 testing sites are distributed unevenly in the US geography and population.
2. Median county-level travel time to SARS-CoV-2 testing sites is longer in less densely populated areas, and in areas with a higher percentage of minority or uninsured populations.
3. Improved geographic accessibility to testing sites is imperative to manage the COVID-19 pandemic in the United States.

## Introduction

Uniform access to SARS-CoV-2 testing is crucial for controlling and containing the COVID-19 epidemic^1^. Lack of testing can result in the epidemic spreading undetected^2,3^ and increase the risk of extensive local transmission. The United States (US) has been slow to develop reliable diagnostic tests and, while there has been recent improvement in testing capabilities^4^, the ability for large-scale testing remains a serious concern.

Inequalities in geographic accessibility to healthcare in the US have been documented to cause negative health outcomes for seasonal influenza transmission and other diseases^5^. Further, travel time negatively impacts healthcare-seeking behavior^6^. The deployment of SARS-CoV-2 testing within existing medical infrastructure, while logistically efficient, may exacerbate this disparity in health outcomes^7,8^ and underestimate disease burden in disadvantaged populations.

Geographic accessibility to SARS-CoV-2 testing sites, to our knowledge, has not been systematically quantified. Therefore, we evaluated whether testing sites were equally accessible to populations across the US, leveraging county-level sociodemographic data, two public SARS-CoV-2 testing site datasets, and a high-resolution map of US travel times.

## Materials and Methods

### Source Population and Covariates

This study was conducted in the 48 contiguous US states and the District of Columbia (DC), using 2014-2018 cross-sectional American Community Survey data (API accessed April 8, 2020, https://www.census.gov/data/developers/data-sets/acs-5year/2018.html), tabulated at the county level. Covariates include population, population density 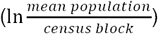, percent uninsured, percent minority (1 − *percent Non Hispanic White*), and median household income.

### Travel Time to Testing Site

#### Testing Sites

A national database of SARS-CoV-2 testing sites was curated using the Carbon Health (N = 5,376) and CodersAgainstCovid (N = 1,547) datasets which were accessed on April 7, 2020. Carbon Health (carbonhealth.com/covid-19-testing-centers) prospectively called urgent care centers and hospitals on publicly listed telephone numbers from March 17, 2020 to ask whether SARS-CoV-19 testing was being offered. Additionally, a verified, non-exhaustive collection of publicly-documented and user-entered testing sites were included. CodersAgainstCovid identified urgent care centers, hospitals, drive-throughs, health departments and other facility types prospectively from March 15, 2020 onward, through volunteer-verified “webscraping” and crowdsourcing (see https://codersagainstcovid.org/).

#### Database compilation and completeness

Testing sites were geocoded using *ggmap* v3.0.0 in R v.3.6.2. We identified 6,236 unique sites (687 excluded following manual de-duplication and cleaning, or because situated in Alaska or Hawaii). Related site ontologies were collapsed into meta-ontologies (e.g. Urgent with Immediate Care). To date, this is the largest database of US testing sites known to the authors. To evaluate completeness (as of April 20, 2020), we identified public testing sites listed in sample areas: 34 in Illinois (https://www.dph.illinois.gov/covid19/covid-19-testing-sites), 5 in Colorado (https://covid19.colorado.gov/testing-covid-19) and 104 in West Virginia (https://www.wvhealthconnection.com/covid-19). Our database included 169, 85 and 60 sites in each area, respectively. We confirmed our database identified at least one site in every city in Texas operating a drive-through (https://www.dshs.state.tx.us/coronavirus/testing.aspx).

#### Travel Time Estimation

We used published friction-based travel times^9^ between approximately 1 km^2^ gridded cells in the US, accounting for topographic features (e.g. water) and the most efficient method of non-air travel (e.g. vehicle). The shortest path to testing sites was calculated using the Dijkstra algorithm^10^. Median travel times across all grid cells in each of the 3,108 counties were calculated.

### Statistical Analysis

Generalized Linear Models (GLM, using *glm, stats* v3.6.2 in R) were used to estimate the effect of population density, percent minority, percent uninsured and median income on median travel time, by county. We also tested for potential interactions between population density and percent minority or percent uninsured. For each model, influential counties with a Cook’s distance measure over 4/*N* were excluded (up to N=175)^11^.

## Results

In our database, we collate 6,236 SARS-Cov-2 testing sites in the 48 contiguous US states plus DC. Testing sites (**Table 1**) were often affiliated with Medical Centers (43%) and Urgent Care (47%). Drive-through sites accounted for only 3% of testing centers. Testing sites were spatially clustered (Moran’s *I*=0.037, *z*=61.4, *p*<10^−5^), likely around US urban centers (**Figure S1)**.

**Table 1.** Breakdown of SARS-CoV-2 Testing Sites by Location Type and Region. Percentage of each type of testing site for the Census Division in parenthesis (48 contiguous US and DC, excluding Hawaii and Alaska). The final column is the total number of testing sites (percentage contributed by that Census Division in parenthesis).

|  | Medical Center<br>N = 2,678 | Urgent Care<br>N = 2,939 | Drive-through<br>N = 187 | Other/Unclassified<br>N = 432 | Total<br>N = 6,236 |
| --- | --- | --- | --- | --- | --- |
| **Geographic Region** |  |  |  |  |  |
| Pacific | 365 (49%) | 290 (39%) | 10 (1%) | 76 (10%) | 741 (12%) |
| Mountain | 261 (47%) | 236 (42%) | 28 (5%) | 31 (6%) | 556 (9%) |
| West North Central | 401 (65%) | 193 (31%) | 5 (1%) | 18 (3%) | 617 (10%) |
| West South Central | 348 (43%) | 377 (46%) | 19 (2%) | 70 (9%) | 814 (13%) |
| East North Central | 434 (55%) | 298 (38%) | 24 (3%) | 27 (3%) | 783 (13%) |
| East South Central | 221 (36%) | 317 (51%) | 14 (2%) | 69 (11%) | 621 (10%) |
| New England | 110 (47%) | 90 (38%) | 23 (10%) | 11 (5%) | 234 (4%) |
| Middle Atlantic | 255 (28%) | 547 (61%) | 20 (2%) | 74 (8%) | 896 (14%) |
| South Atlantic | 283 (29%) | 591 (61%) | 44 (5%) | 56 (6%) | 974 (16%) |

### Spatially Heterogeneous Access to Testing Sites

The travel time from each 1 km^2^ grid cell to the nearest US testing site is spatially heterogeneous at the national and state level (**Figure 1A and 1B**). Thirty percent of the population live in a county (N = 1,920) with a median travel time over 20 minutes, though with pronounced regional differences (**Figure 1C**) ranging from 5% (Middle Atlantic) to 86% (Mountain).

**Figure 1.**
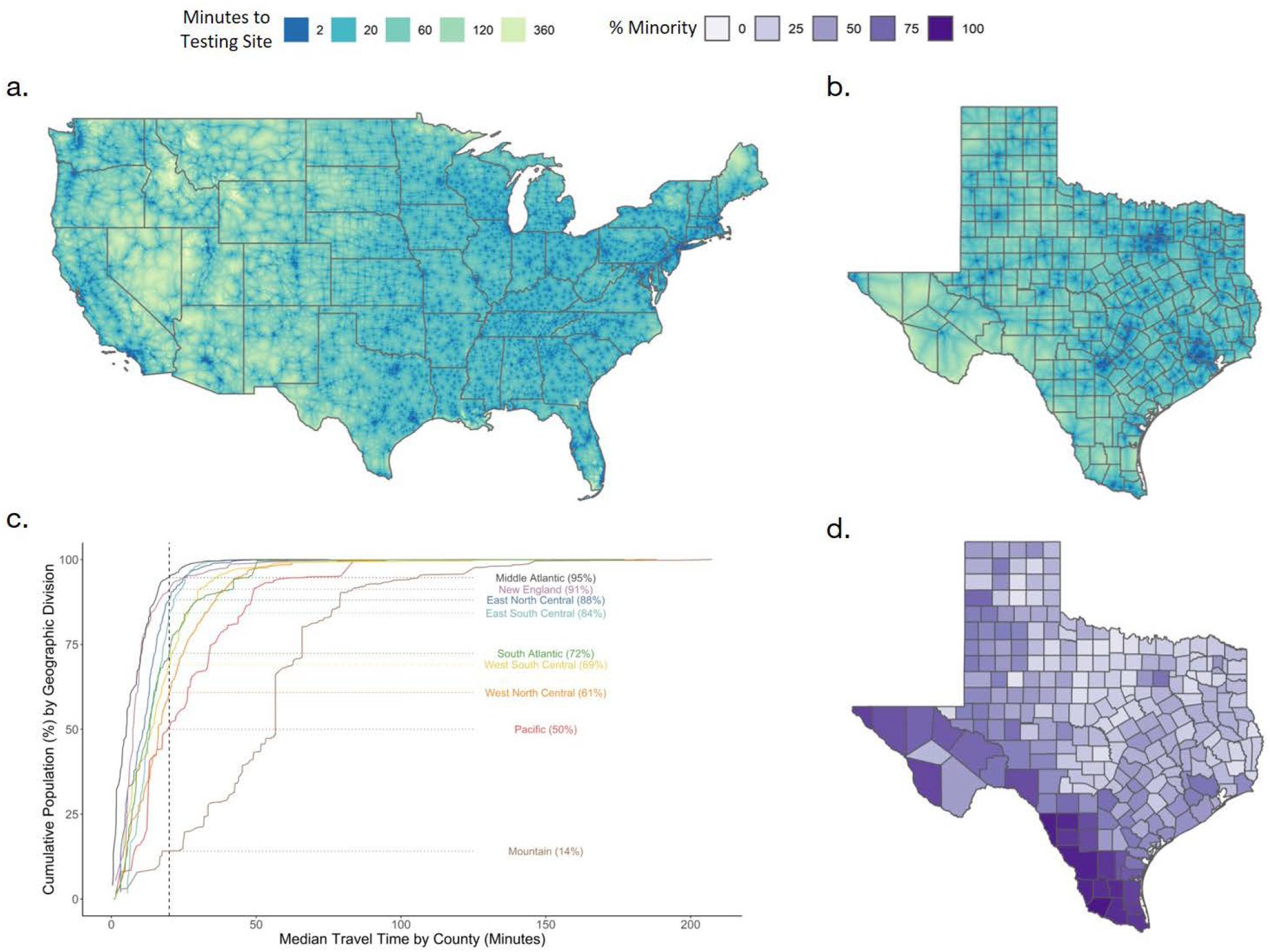
Distribution of SARS-CoV-2 testing sites. A) Travel time to the nearest testing site per 1 km^2^ area (shorter travel time in darker blue) in the 48 contiguous US states plus DC. B) Travel time as in Panel A enlarged to show detail in the state of Texas. C) Median travel time by county versus the cumulative population for each geographic region (excluding two outlier counties). Vertical line at 20 minute median travel time. Horizontal lines indicate cumulative population percentage in that region (in parenthesis) residing in counties with less than 20 minutes median travel time. D) Percent minority (1 − *percent Non Hispanic White*) by county in Texas.

### Determinants of Access to Testing

We estimated the effect of covariates on county-level median travel time (**Table 2**). Population density, a determinant of population distribution, was associated with a shorter median travel time (*β* = -13.41 min/unit density, 95% Confidence Interval (CI) -14.02 to -12.79). While controlling for population density as a potential confounder, percent minority was associated with an increase in travel time (*β* = 0.15 min/percent, 95% CI 0.12-0.18), as was percent uninsured (*β* = 0.41 min/percent, 95% CI 0.3-0.53). These associations remained when also adjusting for median income. We found a significant negative interaction between percent uninsured and population density (*p*<0.01) suggesting that the disparity of longer rural travel times is greater in counties where a higher proportion of the population is uninsured. Percent minority and population density did not interact statistically.

**Table 2:** Generalized Linear Regression models evaluating the association between covariates and median travel time in minutes by county in the 48 contiguous US states and DC.

|  | Model 1 | Model 2 | Model 3 | Model 4 |
| --- | --- | --- | --- | --- |
| Intercept | 61.45 ***<br>[59.82, 63.08] | 59.99 ***<br>[58.34, 61.64] | 56.29 ***<br>[54.17, 58.41] | 51.36 ***<br>[47.70, 55.03] |
| Log of Population Density | -13.41 ***<br>[-14.02, -12.79] | -14.14 ***<br>[-14.76, -13.52] | -12.94 ***<br>[-13.56, -12.32] | -14.13 ***<br>[-14.78, -13.47] |
| Percent Minority (%) |  | 0.15 ***<br>[0.12, 0.18] |  | 0.13 ***<br>[0.10, 0.17] |
| Percent Uninsured (%) |  |  | 0.41 ***<br>[0.30, 0.53] | 0.23 **<br>[0.09, 0.38] |
| Median Income (\$10,000's) | | | | 2.52 ***<br>[1.46, 3.59] |
| N | 2942 | 2934 | 2942 | 2931 |
| AIC | 24321.32 | 24192.59 | 24291.55 | 24097.61 |
| Pseudo R2 | 0.38 | 0.41 | 0.39 | 0.41 |

\*\*\* p < 0.001; \*\* p < 0.01; \* p < 0.05.

## Discussion

Using two large, national datasets of SARS-CoV-2 testing sites throughout the US paired with geographically precise estimates of travel times, we demonstrate an uneven distribution of critical public health resources. The testing site distribution recapitulates structural disparities, including inequities among minority, uninsured, and rural groups, which may further perpetuate disparities as the pandemic progresses. Differential accessibility to testing may lead to biases in estimation of disease incidence, in turn, potentially delaying identification of COVID-19 hot spots. In the absence of representative testing, syndromic surveillance tools (e.g. Safepaths.mit.edu, CovidNearYou.org) may provide early warning signals, and augment targeted-testing and other public health interventions.

The presented analyses are limited by the databases on which they are based. Despite efforts to ensure comprehensiveness, in some regions our dataset may be missing testing sites (e.g. West Virginia) and a handful of sites have been added to source datasets since accession.

Given recent difficulties scaling up testing, we believe our database remains representative^12,13^. However, there is potential for differential missingness of sites located in areas with reduced “webscraping” visibility or sites specifically placed to address geographic inaccessibility.

Nevertheless, this work highlights the potential utility of data sharing during a pandemic, and the need for comprehensive resources to identify geographic gaps and optimize access for all.

The travel time metric used here accounts for the presence of public transportation and routine traffic. Early evidence shows that there is widespread geographic variability in mobility reductions during the epidemic in the US^14,15^. Our estimates of differential access present a conservative picture of inequality in the continental US which may be worse if public transit closures and private transportation were also modeled, and should be the subject of future research. Additionally, our models do not examine other, non-geographic barriers to SARS-CoV-2 testing access (e.g. economic), nor the unique issues for residents in Alaska and Hawaii.

In summary, reduced geographic access to SARS-CoV-2 testing sites is associated with sociodemographic factors that, in turn, are linked to poor structural access to care and health outcomes. The location of future testing sites should explicitly account for travel time and sociodemographic predictors, in addition to other public health mandates for testing.

## Data Availability

For access to the Carbon Health dataset, please. For access to the CodersAgainstCOVID dataset, visit: github.com/codersagainstcovidorg.

## Acknowledgements

The authors would like to thank Mason M. Astley and Kathryn Cordiano for their thoughtful contributions to the manuscript, and Emily Cohn for research support for this analysis. Testing locations were curated and made open-source by volunteers (https://codersagainstcovid.org/about-us) from CodersAgainstCOVID. Testing site data and consultation was also kindly provided by the CarbonHealth Team (https://carbonhealth.com/coronavirus). BR and JSB acknowledge funding from Google.org for COVID-19 research (Tides Foundation, TF2003-089662). CMA was supported by NIDDK (K23 DK120899) and Boston Children’s Hospital (OFD CDA). MUGK acknowledges funding from the MOOD project and a Branco Weiss Fellowship. The funding bodies had no role in study design, data collection and analysis, preparation of the manuscript, or the decision to publish. All authors have seen and approved the manuscript.

## Author contributions

BR, CMA, JSB, MUGK contributed to conceptualization. BR, KTLS, KS contributed to data acquisition. BR, CMA, KTLS contributed to data analysis. All authors contributed to interpretation of results and manuscript writing.

## Data and Code availability

Mean travel time to testing center by county, sociodemographic variables and code for main analyses are available (LINK: XX). Raster of travel times to testing sites is available openly (LINK: XX). For access to the Carbon Health dataset, please. For access to the CodersAgainstCOVID dataset, visit: github.com/codersagainstcovidorg.

## Supplement

**Figure S1.**
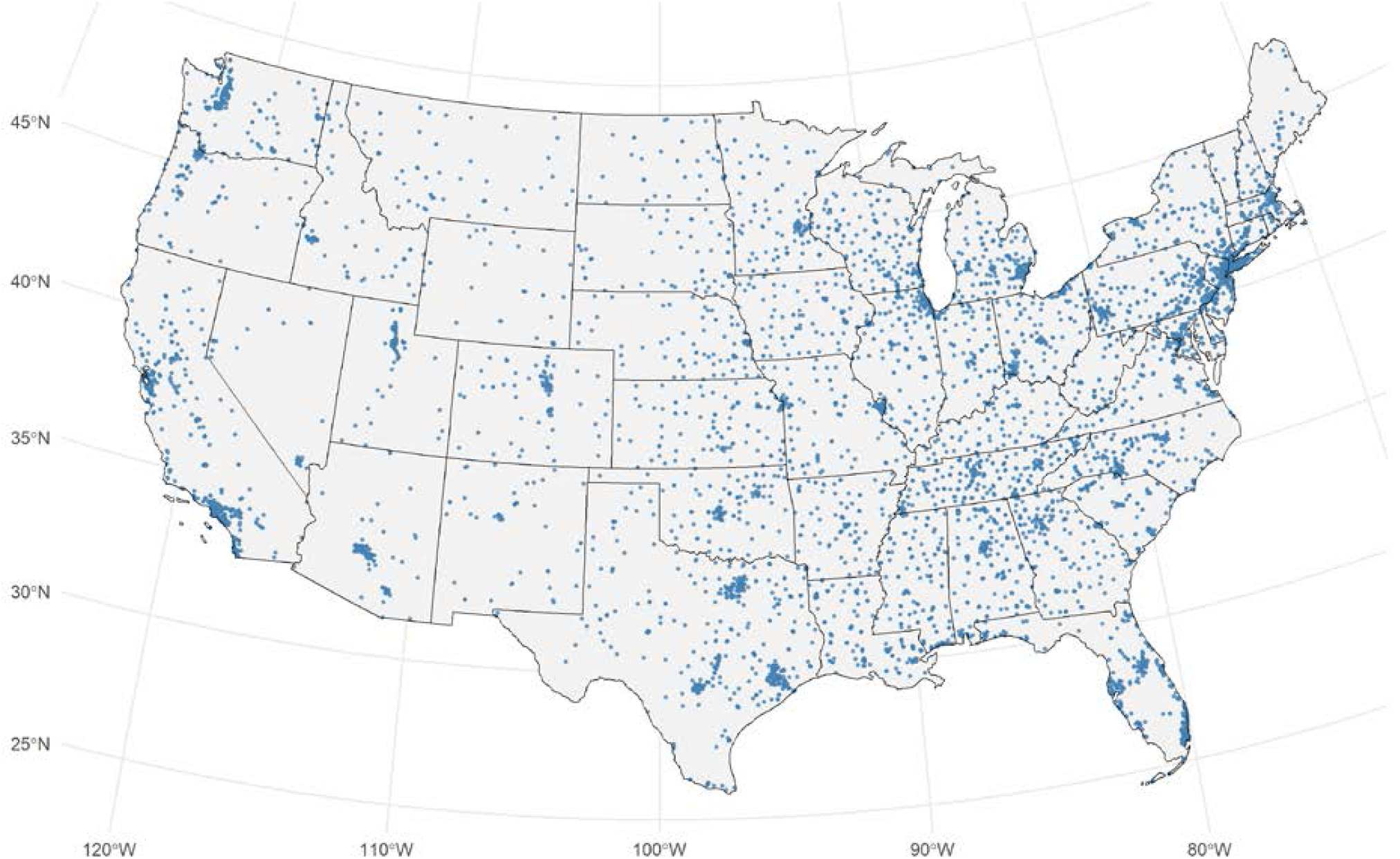
Locations (N = 6,236) offering SARS-CoV-2 testing in the 48 contiguous United States and DC.

